# Distinct neuronal, proliferative, and secretory pathways are perturbed in cancer survivors with depressive symptoms

**DOI:** 10.64898/2026.06.16.26355016

**Authors:** Julia Trudeau, Nidhi Thati, Ding Quan Ng, Esther Chavez-Iglesias, Adam B. Olshen, Anand Dhruva, Jason W. Chan, Raymond J. Chan, Alexandre Chan, Kord M. Kober

## Abstract

**Introduction:** Depression is highly prevalent among cancer survivors and may be biologically distinct, although clinical studies investigating these mechanisms remain limited. Thus, the aims of this study were to (1) identify perturbed biological pathways associated with depressive symptom severity in cancer survivors, and (2) investigate whether these pathways are common or distinct to those perturbed in an age-matched non-cancer cohort.

**Methods:** We analyzed cross-sectional self-reported and transcriptomic data from the Multi-Ethnic Study of Atherosclerosis (PHD #39341). Cancer survivors and an age-matched non-cancer cohort (target ratio 1:2) were identified. The 20-item Center for Epidemiologic Studies Depression Scale (CES-D) was used to split participants into low (CES-D<16) and high (≥16) depressive symptom groups. Analyses were conducted separately for survivor and non-cancer cohorts. Differential gene expression between depressive symptom groups was evaluated with adjustments for covariates significantly associated with depression (survivor cohort: BMI; non-cancer cohort: marital status), with pathway impact analysis identifying perturbed pathways (FDR < 0.025).

**Results:** Ninety-three cancer survivors (11.8% with high depressive symptoms) and 176 non-cancer participants (9.7% with high depressive symptoms) were included. Sixty-eight and 72 perturbed pathways were associated with depression among survivor and non-cancer cohorts, respectively. Twenty-one of these pathways were perturbed uniquely among cancer survivors, which were related to neurodegeneration, reward circuitry, proliferation, and secretion. Inflammatory pathways were consistently perturbed across both cohorts.

**Conclusions:** Distinct biological mechanisms related to neurodegeneration, reward circuitry, autonomic secretion, and proliferative signaling may underlie depression in cancer survivors. Inflammation was implicated as a shared mechanism of depression across cancer and non-cancer populations. This study identifies potential therapeutic targets and highlights the need for precision medicine in treating depression among cancer survivors.

## 3.1 Introduction

Cancer patients experience significantly higher rates of depression relative to the general population, with these symptoms often persisting years into survivorship.^1,2^ While a cancer diagnosis and subsequent treatments are associated with an expected increase in emotional burden, a subset of survivors experience debilitating depressive symptoms that can interfere with their daily quality of life, adherence to survivorship care, and overall outcomes.^3^ Moreover, depression has emerged as a major unmet need in cancer care, with previous studies noting both its under-recognition and undertreatment.^4,5^ For instance, several systematic reviews^6–8^ and the American Society of Clinical Oncology (ASCO) clinical guidelines^9^ largely support non-pharmacologic interventions and highlight a lack of robust evidence supporting the efficacy of commonly used antidepressants in cancer populations. As a result, there is an urgent need for an increased understanding of the mechanisms underlying depression among cancer survivors to inform future targeted treatment strategies.

This is particularly relevant amid emerging hypotheses that the pathophysiology underlying depressive symptoms among cancer patients may be biologically distinct from the general population. Rather than being explained solely as a reactionary outcome to a cancer diagnosis and its associated psychological and physical burdens, such as anxiety and other co-occuring symptoms, growing evidence points to potential population-specific biological dysregulations as contributors to the disproportionately high prevalence of depression.^2,10^ Major biological hypotheses of depression include the traditional monoamine hypothesis, the hypothalamic-pituitary-adrenal (HPA) axis hypothesis, the cytokine hypothesis, and the neuroplasticity hypothesis.^11^ Previous reviews focusing on these mechanisms in the specific context of cancer posit that overlaps in the biological environment of cancer and depression may foster their coexistence.^2,10,12^ For instance, cancer and anti-cancer therapies may increase the release of pro-inflammatory cytokines, which can drive neuroinflammation and subsequent depressive symptoms.^13^ This is supported by a pre-clinical model in which cancer induced inflammation and depressive-like behaviors in mice,^14^ as well as clinical studies showing that elevations of circulating inflammatory cytokines are associated with depressive symptoms in cancer populations.^12,15^ However, these associations have also been observed in non-cancer populations,^16^ and clinical evidence regarding inflammation as a cancer-specific mechanism of depression remains scarce. Furthermore, an overactive HPA axis is characteristic of depression, and it has been proposed that this mechanism may be especially relevant in cancer populations, considering their exposure to chronic stress and cancer-induced inflammation.^2,10^

Despite these emerging hypotheses, evidence of molecular mechanisms underlying depression among cancer survivors, and in particular how they may differ from the general population, remains extremely limited. Data-driven transcriptomic analyses can provide important insights into these mechanisms to help inform investigations into targeted interventions. However, studies evaluating mechanisms of depression using gene expression data from human samples have primarily been conducted using general, rather than cancer-specific, populations.^17–19^ Furthermore, several previous transcriptomic studies identified differentially expressed genes across both cancer conditions and major depressive disorder (MDD) to investigate shared mechanisms, rather than characterizing depression within cancer populations.^20–22^ One exploratory transcriptomic analysis in a small sample (n=10) of ovarian cancer patients identified genes and pathways associated with depression.^23^ However, these findings cannot represent cancer survivors as a whole and do not provide insight on how mechanisms differ relative to non-cancer groups.

Pathway impact analysis allows for the interpretation of differential gene expression data to identify perturbed biological signaling pathways in a clinical phenotype, expanding beyond enrichment analysis by considering relevant factors such as gene-gene interactions and the magnitude of differential expression.^24,25^ Thus, this study utilizes this approach using transcriptomic data from participants in the Multi-Ethnic Study of Atherosclerosis (MESA) study to (1) evaluate for perturbed signaling pathways associated with depressive symptoms among cancer survivors, and (2) contextualize the findings relative to those in age-matched non-cancer participants to evaluate for pathways underlying depression that are uniquely perturbed among cancer survivors. Findings will further our understanding of mechanisms of depression in this population to inform precision medicine approaches in survivorship care.

## 3.2 Methods

### 3.2.1 Participants and Data Source

This is a cross-sectional analysis of gene expression and self-reported data from participants in the Multi-Ethnic Study of Atherosclerosis (MESA), a prospective longitudinal study.^26,27^ The MESA study was designed to recruit a diverse, US population-based sample from six geographically diverse field centers to understand risk factors associated with subclinical cardiovascular disease. Data were collected on a range of sociodemographic, psychosocial, and molecular measures. Eligible participants were between 45 and 84 years old, identified as White, African-American, Chinese-American, or Hispanic, and did not meet any of the exclusion criteria.^27^ Exclusion criteria included active treatment for cancer and serious medical conditions. According to the National Cancer Institute, an individual is considered a cancer survivor from the time of diagnosis through the end of life.^28^ This definition aligns with any MESA participant with a self-report of a history of a cancer diagnosis (i.e., “Has a doctor ever told you that you had any of the following: Cancer”). Gene expression data were available from subsets of participants from Exam 1 (2000-2002) and Exam 5 (2010-2011). For participants with data at both exams, one observation was selected to ensure independence using the decision criteria outlined in Supplementary Figure 1. Data from the MESA study were obtained through an approved study project (39341) and dbGaP studies phs000209.v13.p3 (Multi-Ethnic Study of Atherosclerosis (MESA) Cohort) and phs001416.v4.p1 (NHLBI TOPMed MESA and MESA Family AA-CAC).

### 3.2.2 Instruments

Participants completed a comprehensive set of questionnaires (e.g., medical history, psycho-social), including the 20-item Center for Epidemiologic Studies Depression Scale (CES-D). The CES-D is a validated and widely used scale for screening depression with good psychometrics, including in cancer populations.^29–32^ Participants rank how often they have experienced certain feelings (i.e., “I felt sad”) over the past week on a 4-point Likert scale and items are added to achieve a total score, with higher representing worse depressive symptoms (Supplementary Table 1). While this tool is not intended to diagnose MDD, an established cutoff of ≥16 is used to indicate an individual at risk for clinical depression.^33,34^ As such, we classified participants into low (<16) and high (≥16) depressive symptom groups. Participants with missing CES-D data were excluded.

Sociodemographic (age, gender, race/ethnicity, household income, marital status, body mass index [BMI]) and clinical characteristics (antidepressant use, diabetes, hypertension, cancer diagnosis if identified as survivor) were assessed. Missing data for these characteristics were imputed by the k-nearest-neighbors method, with k=9. For continuous variables, the Euclidean distance was used to find the nearest neighbors.

The imputed value was the weighted average of the nearest neighbors, with each weight originally exp(-dist(x,j)), after which the weights were scaled to one. For categorical variables, distance was 0 if the predictor and the neighbor had the same value and 1 if they did not and the imputed value was the mode of the nearest neighbors. Gene expression was assessed using RNA isolated from peripheral blood mononuclear cells (PBMCs) and quantified as expected counts.^35^

### 3.2.3 Data analysis

#### Cancer survivor and matched non-cancer cohorts

All analyses were completed using R (version 4.1, https://www.R-project.org/). To determine whether findings were distinct to cancer survivors, we created an age-matched non-cancer cohort from MESA study participants who answered “No” to “Has a doctor ever told you that you had any of the following: Cancer” using the R package MatchIt.^36^ Consistent with the categorization schema in the MESA dataset, age was grouped in 10-year intervals and non-cancer survivors were age-matched to cancer survivors using the nearest-neighbor algorithm (caliper width=0.2, target ratio 1 cancer survivor: 2 non-cancer).^37,38^ Successful matching was defined as all age groups having standardized mean differences of <|0.1|.^39^ Post-matching differences in sociodemographic characteristics between cohorts were evaluated using chi-square and Fisher’s exact tests as appropriate.

#### Patient demographic, clinical, and psychosocial data

All further analyses were performed separately for cancer and non-cancer cohorts. Differences in sociodemographic and clinical characteristics between low and high depressive groups were evaluated using chi-square and Fisher’s exact tests as appropriate.

#### Differential gene expression

Following our previous studies, differential gene expression between low and high depressive symptom groups was quantified using empirical Bayes models in edgeR.^25,40–43^ Sociodemographic and clinical characteristics that significantly differed (p<0.05) between low and high depressive symptom groups were included as covariates. Surrogate variables not associated with depression were included to adjust for variation due to unmeasured sources.^44^

#### Pathway impact analysis (PIA)

Differential expression results were interpreted using Pathway Impact Analysis (PIA) to identify perturbed pathways and provide insight on depression-related mechanisms.^43^ PIA interprets the results of the differential expression analyses for all genes (i.e., cutoff-free) to determine the probability of pathway perturbations using Pathway Express.^45^ A total of 149 signaling pathways were identified using the Kyoto Encyclopedia of Genes and Genomes (KEGG) database (Release 73.0+/01-03, Jan 15).^46^ For each cohort, a separate test was performed for each pathway. Significance for the transcriptome-wide PIA was determined using a strict false discovery rate (FDR) of 0.025 under the Benjamini-Hochberg procedure.^47^ Pathways associated with depressive symptoms were evaluated separately in cancer survivors and age-matched non-cancer cohorts to identify pathways that were common across both groups and those potentially distinct to cancer survivors.

## 3.3 Results

### 3.3.1 Participant characteristics

93 cancer survivors (n=11 [11.8%] with high depressive symptoms) and 176 non-cancer participants (n=17 [9.7%] with high depressive symptoms) were included. Cancer survivors were predominantly 75-84 years old (51%), female (52%), non-Hispanic White (67%), and most had other, multiple, or unspecified prior cancer diagnoses (44%) (Table 1). Age-matching was successful, with all standardized mean differences <|0.1| (Figure 1). Pre-matched non-cancer participants were significantly younger than cancer survivors (p<0.001), with no differences in the matched cohort (p=0.975) (Supplementary Table 2). Differences remained between cancer survivors and the age-matched non-cancer cohort in race/ethnicity and household income (p<0.05) (Supplementary Table 2). Among cancer survivors, although there was an association indicated between depression with BMI (*x*^2^=10.32; p=0.018) (Table 1), post-hoc comparisons using Fisher’s exact test with Bonferroni correction did not show statistically significant differences between individual BMI categories, though there was a trend toward higher depressive symptoms in more severely overweight (Grade 3) survivors compared with moderately overweight (Grade 2) survivors (p=0.023, adjusted p=0.14). Among the non-cancer cohort, those who were unmarried were more likely to have high depressive symptoms (OR=0.24; p=0.019) (Table 2). BMI and marital status were subsequently included in differential gene expression analyses in the cancer and non-cancer cohorts, respectively.

**Figure 1.**
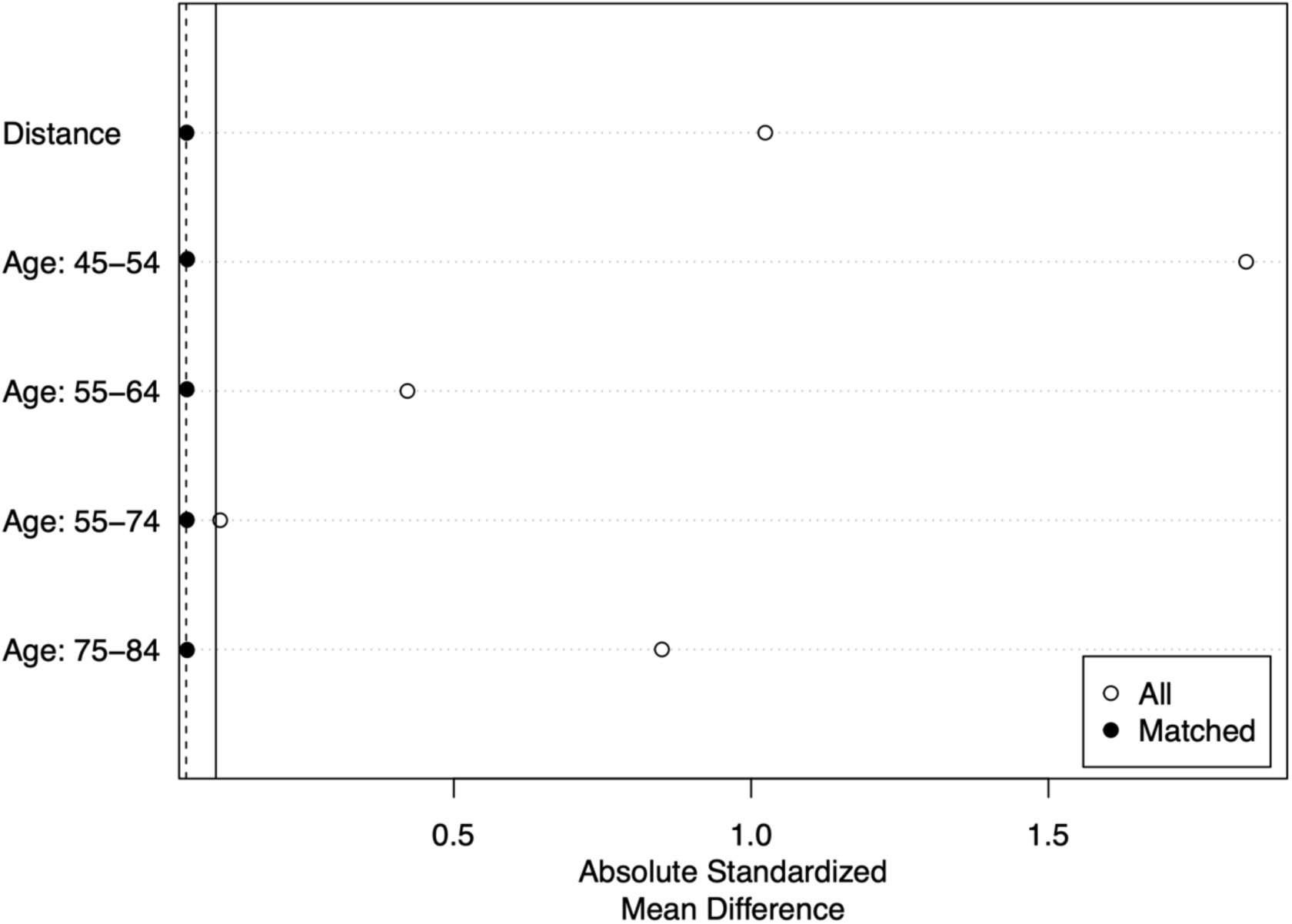
Love plot depicting absolute standardized mean differences in age groups between cancer and non-cancer cohorts before and after age-matching.

**Table 1.**
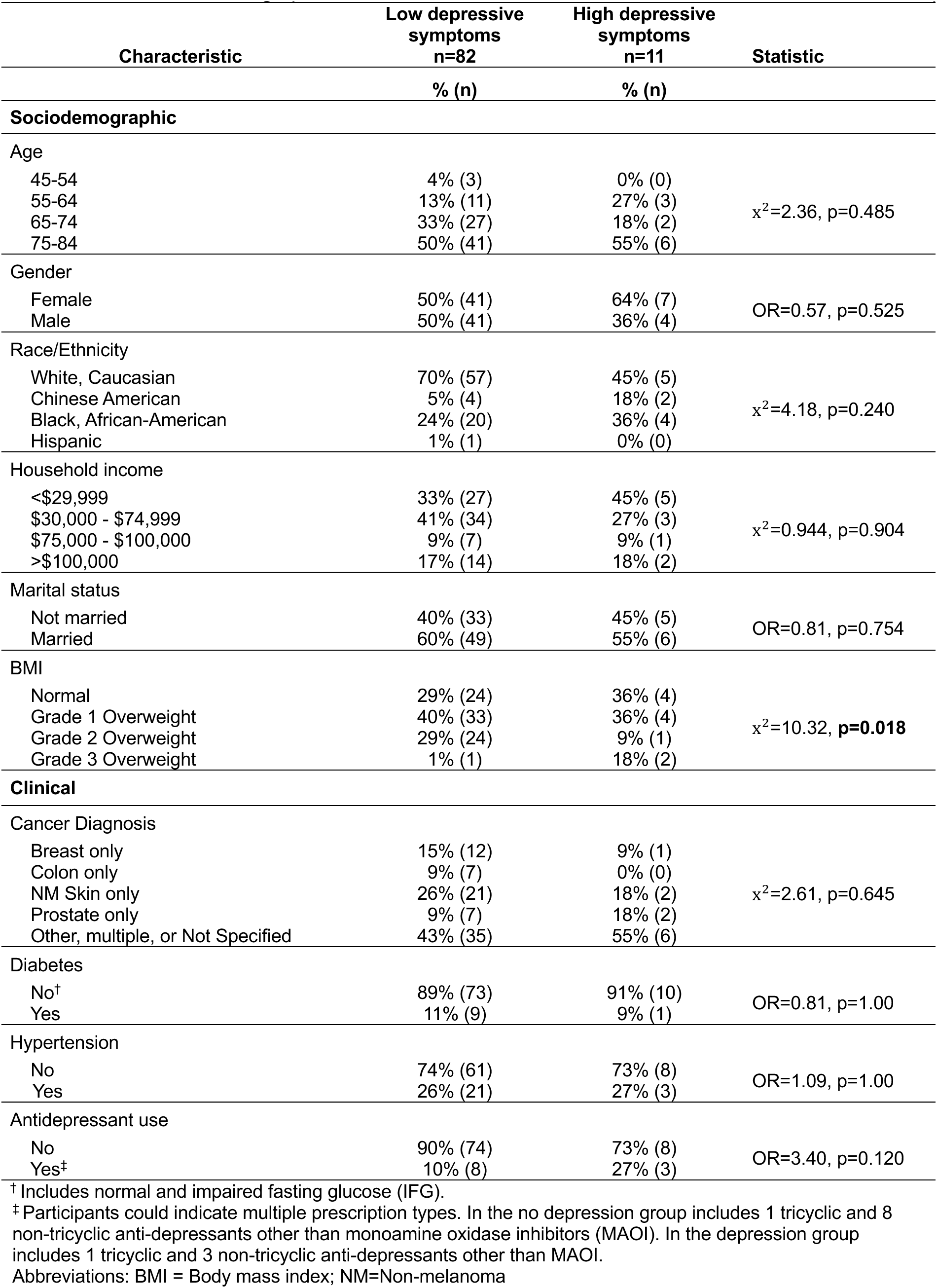
Differences in demographic and clinical characteristics in cancer survivors with and without depression.

**Table 2.** Differences in demographic and clinical characteristics in age-matched non-cancer cohort with and without depression.

| Characteristic | Low depressive symptoms<br>n=159 | High depressive symptoms<br>n=17 | Statistic |
| --- | --- | --- | --- |
|  | % (n) | % (n) |  |
| Sociodemographic |  |  |  |
| Age |  |  |  |
| 45-54 | 3% (5) | 6% (1) | x<sup>2</sup>=1.37, p=0.745 |
| 55-64 | 16% (25) | 18% (3) |  |
| 65-74 | 32% (51) | 41% (7) |  |
| 75-84 | 49% (78) | 35% (6) |  |
| Gender |  |  |  |
| Female | 55% (88) | 71% (12) | OR=0.52, p=0.305 |
| Male | 45% (71) | 29% (5) |  |
| Race/Ethnicity |  |  |  |
| White, Caucasian | 35% (55) | 24% (4) | x<sup>2</sup>=6.33, p=0.100 |
| Chinese American | 15% (24) | 0% (0) |  |
| Black, African-American | 30% (48) | 35% (6) |  |
| Hispanic | 20% (32) | 41% (7) |  |
| Household income |  |  |  |
| <\$29,999 | 49% (78) | 59% (10) | x<sup>2</sup>=1.13, p=0.783 |
| \$30,000 - \$74,999 | 38% (61) | 35% (6) | |
| \$75,000 - \$100,000 | 4% (6) | 0% (0) | |
| >\$100,000 | 9% (14) | 6% (1) | |
| Marital status |  |  |  |
| Not married | 44% (70) | 77% (13) | OR=0.24, **p=0.019** |
| Married | 56% (89) | 24% (4) |  |
| BMI |  |  |  |
| Normal | 29% (46) | 29% (5) | x<sup>2</sup>=0.20, p=1.00 |
| Grade 1 Overweight | 38% (61) | 35% (6) |  |
| Grade 2 Overweight | 32% (51) | 35% (6) |  |
| Grade 3 Overweight | 1% (1) | 0% (0) |  |
| Clinical |  |  |  |
| Diabetes |  |  |  |
| No<sup>†</sup> | 88% (140) | 88% (15) | OR=0.98, p=1.00 |
| Yes | 12% (19) | 12% (2) |  |
| Hypertension |  |  |  |
| No | 64% (102) | 65% (11) | OR=0.98, p=1.00 |
| Yes | 36% (57) | 35% (6) |  |
| Antidepressant use |  |  |  |
| No | 95% (152) | 94% (16) | OR=1.35, p=0.564 |
| Yes<sup>‡</sup> | 4% (7) | 6% (1) |  |

<sup>†</sup> Includes normal and impaired fasting glucose (IFG).
<sup>‡</sup> Participants could indicate multiple prescription types. In the no depression group includes 2 tricyclic and 5 non-tricyclic anti-depressants other than monoamine oxidase inhibitors (MAOI). In the depression group all were non-tricyclic anti-depressants other than MAOI.
Abbreviations: BMI = Body mass index

### 3.3.2 Differential expression and pathway perturbation

Differential expression results are available as supplemental files. Of the 68 pathways that were significantly perturbed between low and high depressive symptom groups among cancer survivors (Tables 3-5, Supplemental File 1), 21 of these pathways were perturbed only among cancer survivors. A total of 72 pathways were significantly perturbed between low and high depressive symptom groups in the age-matched non-cancer cohort (Tables 3-5, Supplemental File 2). Perturbed pathways across both cohorts were related to (1) neuronal functioning and neurotransmission, (2) cell cycle, growth, and proliferation, (3) cell adhesion and cytoskeleton, (4) metabolism and endocrine signaling, (5) cardiovascular processes, and (6) inflammatory processes and immune signaling.

**Table 3.** Perturbed KEGG signaling pathways related to neuronal functioning, neurotransmission, and reward circuitry associated with depression.

|  |  | Cancer Survivors |  | Non-Cancer Controls |  |
| --- | --- | --- | --- | --- | --- |
| Pathway ID | Pathway Name | Pert. Score<sup>†</sup> | FDR<sup>‡</sup> | Pert. Score<sup>†</sup> | FDR<sup>‡</sup> |
| Neuronal Functioning, Neurotransmission, and Reward Circuitry |  |  |  |  |  |
| Common |  |  |  |  |  |
| hsa04080 | Neuroactive ligand-receptor interaction | 12.56 | 0.006 | 9.93 | 0.002 |
| hsa05020 | Prion diseases | 17.88 | 0.006 | 17.33 | 0.002 |
| hsa04020 | Calcium signaling pathway | 4.81 | 0.010 | 3.70 | 0.020 |
| hsa04360 | Axon guidance | 7.68 | 0.011 | 9.26 | 0.002 |
| hsa04740 | Olfactory transduction | 4.87 | 0.015 | 4.24 | 0.012 |
| hsa04723 | Retrograde endocannabinoid signaling | 3.14 | 0.024 | 7.06 | 0.003 |
| Distinct to Cancer Survivors |  |  |  |  |  |
| hsa05016 | Huntington's disease | 3.98 | 0.013 | -0.95 | 0.399 |
| hsa05031 | Amphetamine addiction | 6.02 | 0.013 | 1.95 | 0.079 |
| hsa05034 | Alcoholism | 5.67 | 0.014 | 1.98 | 0.075 |
| hsa04721 | Synaptic vesicle cycle | 4.13 | 0.015 | 1.31 | 0.202 |
| hsa05014 | Amyotrophic lateral sclerosis (ALS) | 4.25 | 0.017 | 2.96 | 0.039 |
| hsa05010 | Alzheimer's disease | 3.89 | 0.018 | 2.70 | 0.037 |
| hsa04713 | Circadian entrainment | 4.14 | 0.021 | 0.86 | 0.414 |
| hsa04744 | Phototransduction | 3.82 | 0.021 | 0.48 | 0.597 |
| hsa05030 | Cocaine addiction | 3.77 | 0.021 | 1.36 | 0.133 |
| hsa04728 | Dopaminergic synapse | 3.36 | 0.024 | 0.78 | 0.473 |
| Distinct to Non-Cancer Controls |  |  |  |  |  |
| hsa04912 | GnRH signaling pathway | 2.28 | 0.054 | 3.59 | 0.021 |
| hsa04726 | Serotonergic synapse | 0.91 | 0.318 | 7.74 | 0.003 |

<sup>†</sup> Normalized pathway perturbation scores. Higher values represent greater pathway perturbation.
<sup>‡</sup> Bold indicates significance at a false discovery rate (FDR) of < 0.025.
Abbreviations: KEGG, Kyoto Encyclopedia of Genes and Genomes.

**Table 4.**
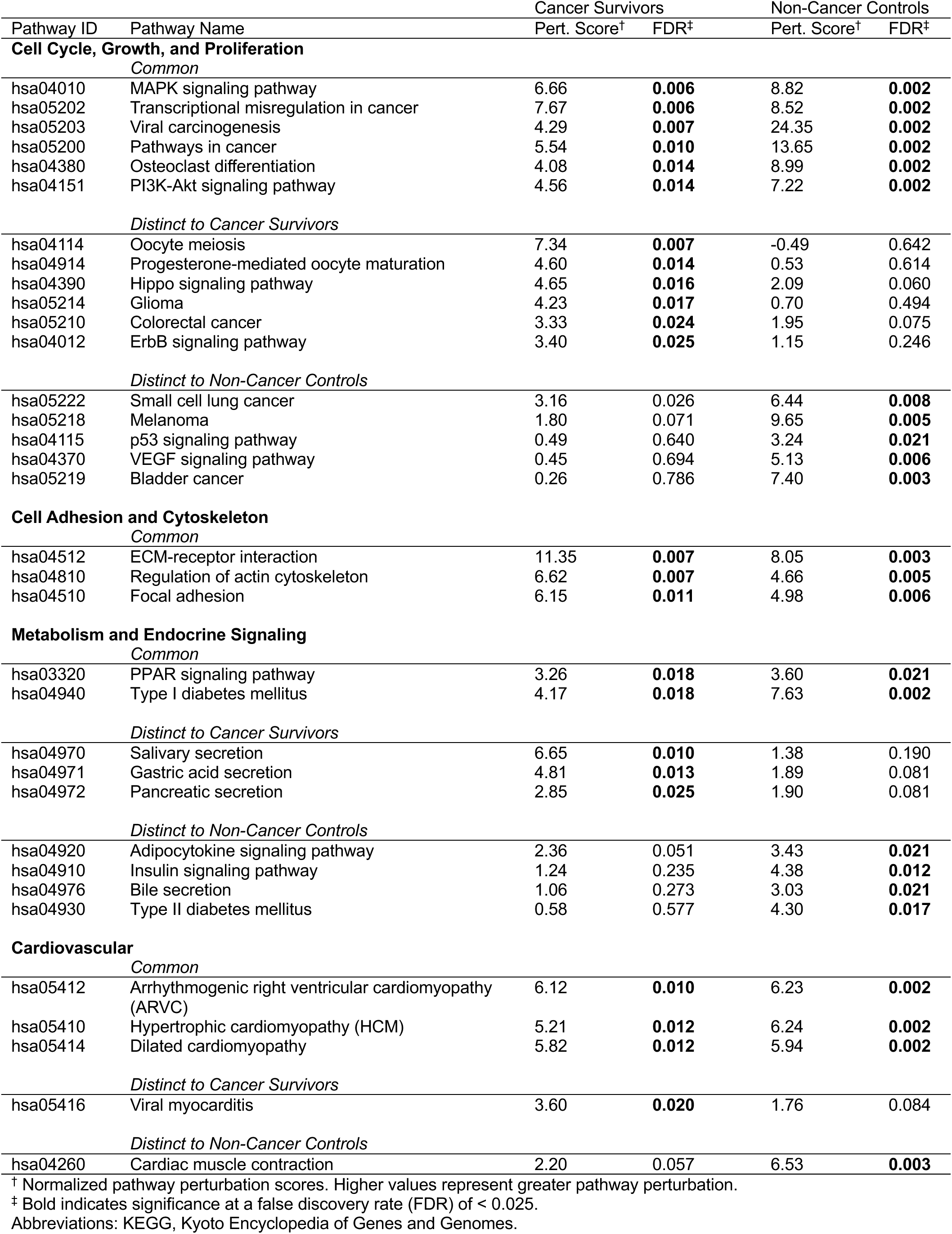
Perturbed KEGG signaling pathways related to cell cycle, growth, and proliferation, cell adhesion and cytoskeleton, and metabolism and endocrine signaling associated with depression.

|  |  | Cancer Survivors |  | Non-Cancer Controls |  |
| --- | --- | --- | --- | --- | --- |
| Pathway ID | Pathway Name | Pert. Score<sup>†</sup> | FDR<sup>‡</sup> | Pert. Score<sup>†</sup> | FDR<sup>‡</sup> |
| Cell Cycle, Growth, and Proliferation |  |  |  |  |  |
| Common |  |  |  |  |  |
| hsa04010 | MAPK signaling pathway | 6.66 | 0.006 | 8.82 | 0.002 |
| hsa05202 | Transcriptional misregulation in cancer | 7.67 | 0.006 | 8.52 | 0.002 |
| hsa05203 | Viral carcinogenesis | 4.29 | 0.007 | 24.35 | 0.002 |
| hsa05200 | Pathways in cancer | 5.54 | 0.010 | 13.65 | 0.002 |
| hsa04380 | Osteoclast differentiation | 4.08 | 0.014 | 8.99 | 0.002 |
| hsa04151 | PI3K-Akt signaling pathway | 4.56 | 0.014 | 7.22 | 0.002 |
| Distinct to Cancer Survivors |  |  |  |  |  |
| hsa04114 | Oocyte meiosis | 7.34 | 0.007 | -0.49 | 0.642 |
| hsa04914 | Progesterone-mediated oocyte maturation | 4.60 | 0.014 | 0.53 | 0.614 |
| hsa04390 | Hippo signaling pathway | 4.65 | 0.016 | 2.09 | 0.060 |
| hsa05214 | Glioma | 4.23 | 0.017 | 0.70 | 0.494 |
| hsa05210 | Colorectal cancer | 3.33 | 0.024 | 1.95 | 0.075 |
| hsa04012 | ErbB signaling pathway | 3.40 | 0.025 | 1.15 | 0.246 |
| Distinct to Non-Cancer Controls |  |  |  |  |  |
| hsa05222 | Small cell lung cancer | 3.16 | 0.026 | 6.44 | 0.008 |
| hsa05218 | Melanoma | 1.80 | 0.071 | 9.65 | 0.005 |
| hsa04115 | p53 signaling pathway | 0.49 | 0.640 | 3.24 | 0.021 |
| hsa04370 | VEGF signaling pathway | 0.45 | 0.694 | 5.13 | 0.006 |
| hsa05219 | Bladder cancer | 0.26 | 0.786 | 7.40 | 0.003 |
| Cell Adhesion and Cytoskeleton |  |  |  |  |  |
| Common |  |  |  |  |  |
| hsa04512 | ECM-receptor interaction | 11.35 | 0.007 | 8.05 | 0.003 |
| hsa04810 | Regulation of actin cytoskeleton | 6.62 | 0.007 | 4.66 | 0.005 |
| hsa04510 | Focal adhesion | 6.15 | 0.011 | 4.98 | 0.006 |
| Metabolism and Endocrine Signaling |  |  |  |  |  |
| Common |  |  |  |  |  |
| hsa03320 | PPAR signaling pathway | 3.26 | 0.018 | 3.60 | 0.021 |
| hsa04940 | Type I diabetes mellitus | 4.17 | 0.018 | 7.63 | 0.002 |
| Distinct to Cancer Survivors |  |  |  |  |  |
| hsa04970 | Salivary secretion | 6.65 | 0.010 | 1.38 | 0.190 |
| hsa04971 | Gastric acid secretion | 4.81 | 0.013 | 1.89 | 0.081 |
| hsa04972 | Pancreatic secretion | 2.85 | 0.025 | 1.90 | 0.081 |
| Distinct to Non-Cancer Controls |  |  |  |  |  |
| hsa04920 | Adipocytokine signaling pathway | 2.36 | 0.051 | 3.43 | 0.021 |
| hsa04910 | Insulin signaling pathway | 1.24 | 0.235 | 4.38 | 0.012 |
| hsa04976 | Bile secretion | 1.06 | 0.273 | 3.03 | 0.021 |
| hsa04930 | Type II diabetes mellitus | 0.58 | 0.577 | 4.30 | 0.017 |
| Cardiovascular |  |  |  |  |  |
| Common |  |  |  |  |  |
| hsa05412 | Arrhythmogenic right ventricular cardiomyopathy (ARVC) | 6.12 | 0.010 | 6.23 | 0.002 |
| hsa05410 | Hypertrophic cardiomyopathy (HCM) | 5.21 | 0.012 | 6.24 | 0.002 |
| hsa05414 | Dilated cardiomyopathy | 5.82 | 0.012 | 5.94 | 0.002 |
| Distinct to Cancer Survivors |  |  |  |  |  |
| hsa05416 | Viral myocarditis | 3.60 | 0.020 | 1.76 | 0.084 |
| Distinct to Non-Cancer Controls |  |  |  |  |  |
| hsa04260 | Cardiac muscle contraction | 2.20 | 0.057 | 6.53 | 0.003 |

<sup>†</sup> Normalized pathway perturbation scores. Higher values represent greater pathway perturbation.
<sup>‡</sup> Bold indicates significance at a false discovery rate (FDR) of < 0.025.
Abbreviations: KEGG, Kyoto Encyclopedia of Genes and Genomes.

**Table 5.** Perturbed KEGG signaling pathways related to inflammatory processes and immune signaling associated with depression.

|  |  | Cancer Survivors |  | Non-Cancer Controls |  |
| --- | --- | --- | --- | --- | --- |
| Pathway ID | Pathway Name | Pert. Score<sup>†</sup> | FDR<sup>‡</sup> | Pert. Score<sup>†</sup> | FDR<sup>‡</sup> |
| Inflammatory Processes and Immune Signaling |  |  |  |  |  |
| Common |  |  |  |  |  |
| hsa04060 | Cytokine-cytokine receptor interaction | 14.75 | 0.006 | 20.20 | 0.002 |
| hsa04062 | Chemokine signaling pathway | 13.75 | 0.006 | 12.70 | 0.002 |
| hsa05132 | Salmonella infection | 8.92 | 0.006 | 10.41 | 0.003 |
| hsa05134 | Legionellosis | 8.28 | 0.006 | 9.55 | 0.002 |
| hsa05144 | Malaria | 8.29 | 0.006 | 19.90 | 0.002 |
| hsa05146 | Amoebiasis | 12.36 | 0.006 | 10.00 | 0.002 |
| hsa05166 | HTLV-I infection | 8.87 | 0.006 | 21.05 | 0.002 |
| hsa05322 | Systemic lupus erythematosus | 12.11 | 0.006 | 11.57 | 0.002 |
| hsa05323 | Rheumatoid arthritis | 8.84 | 0.006 | 14.03 | 0.002 |
| hsa05133 | Pertussis | 8.11 | 0.007 | 6.26 | 0.002 |
| hsa05142 | Chagas disease (American trypanosomiasis) | 7.43 | 0.007 | 11.34 | 0.002 |
| hsa05150 | Staphylococcus aureus infection | 9.67 | 0.007 | 9.40 | 0.002 |
| hsa05100 | Bacterial invasion of epithelial cells | 7.02 | 0.010 | 3.58 | 0.023 |
| hsa04620 | Toll-like receptor signaling pathway | 5.06 | 0.010 | 7.94 | 0.003 |
| hsa05152 | Tuberculosis | 5.44 | 0.010 | 7.77 | 0.002 |
| hsa05161 | Hepatitis B | 5.16 | 0.010 | 24.38 | 0.002 |
| hsa04145 | Phagosome | 3.87 | 0.011 | 8.78 | 0.002 |
| hsa04610 | Complement and coagulation cascades | 7.70 | 0.011 | 5.12 | 0.011 |
| hsa04612 | Antigen processing and presentation | 4.99 | 0.011 | 8.21 | 0.002 |
| hsa05140 | Leishmaniasis | 4.40 | 0.011 | 11.15 | 0.003 |
| hsa04621 | NOD-like receptor signaling pathway | 5.55 | 0.012 | 12.30 | 0.002 |
| hsa04144 | Endocytosis | 3.34 | 0.013 | 2.86 | 0.023 |
| hsa04064 | NF-kappa B signaling pathway | 4.50 | 0.014 | 9.61 | 0.002 |
| hsa05120 | Epithelial cell signaling in Helicobacter pylori infection | 4.28 | 0.015 | 10.06 | 0.003 |
| hsa05164 | Influenza A | 3.60 | 0.017 | 8.55 | 0.002 |
| hsa05143 | African trypanosomiasis | 5.06 | 0.017 | 15.06 | 0.002 |
| hsa05330 | Allograft rejection | 3.37 | 0.024 | 5.53 | 0.003 |
| Distinct to Cancer Survivors |  |  |  |  |  |
| hsa04666 | Fc gamma R-mediated phagocytosis | 3.37 | 0.015 | 1.08 | 0.261 |
| Distinct to Non-Cancer Controls |  |  |  |  |  |
| hsa05332 | Graft-versus-host disease | 3.50 | 0.026 | 7.74 | 0.002 |
| hsa05310 | Asthma | 2.85 | 0.029 | 4.89 | 0.007 |
| hsa04210 | Apoptosis | 2.87 | 0.035 | 4.42 | 0.017 |
| hsa05162 | Measles | 2.58 | 0.038 | 5.33 | 0.005 |
| hsa05320 | Autoimmune thyroid disease | 2.28 | 0.046 | 3.30 | 0.023 |
| hsa05168 | Herpes simplex infection | 2.15 | 0.054 | 5.09 | 0.007 |
| hsa04622 | RIG-I-like receptor signaling pathway | 2.13 | 0.054 | 5.82 | 0.006 |
| hsa05145 | Toxoplasmosis | 1.99 | 0.065 | 3.57 | 0.017 |
| hsa04650 | Natural killer cell mediated cytotoxicity | 1.73 | 0.090 | 4.30 | 0.012 |
| hsa05160 | Hepatitis C | 1.57 | 0.118 | 4.94 | 0.008 |
| hsa05169 | Epstein-Barr virus infection | 1.33 | 0.166 | 3.50 | 0.017 |
| hsa04630 | Jak-STAT signaling pathway | 1.08 | 0.282 | 4.34 | 0.008 |
| hsa05131 | Shigellosis | 0.91 | 0.344 | 4.81 | 0.012 |

<sup>†</sup> Normalized pathway perturbation scores. Higher values represent greater pathway perturbation.
<sup>‡</sup> Bold indicates significance at a false discovery rate (FDR) of < 0.025.
Abbreviations: KEGG, Kyoto Encyclopedia of Genes and Genomes.

## 3.4 Discussion

This age-matched study utilizes a novel strategy to investigate biological mechanisms of depression among cancer survivors, identifying perturbed pathways within both survivor and non-cancer cohorts. Our findings implicate that depressive symptoms among cancer survivors are associated with perturbations in (1) neuronal functioning, neurotransmission, and reward circuitry (2) cell growth and proliferation, (3) cell adhesion and cytoskeleton, (4) metabolism and endocrine signaling, (5) cardiovascular processes, and (6) inflammation. In all mechanistic categories, excluding cell adhesion and cytoskeleton, we identified specific pathways associated with depressive symptomology that were unique to cancer survivors. Notably, these were related to neurodegeneration, reward circuitry, autonomic secretion, and distinct cell cycle and proliferative processes, with a recurring theme of pathways impacted by biological and psychological stressors such as chronic psychological distress and treatment exposure. These findings provide essential clinical, transcriptomic-wide evidence supporting emerging hypotheses that depression among cancer populations may be biologically distinct and require precision medicine approaches.^2,20,48^ In contrast, while inflammation is frequently cited as a hypothesized mechanism of cancer-associated depression,^2,10,49^ we observed substantial overlap in inflammatory pathway perturbations across both cancer survivor and non-cancer cohorts, suggesting that this may represent a common mechanism across populations.

As social and environmental factors are known to interact with underlying biology to influence depression, we will first discuss our findings regarding demographic and clinical characteristics. We will then discuss perturbations within each mechanistic category with a focus on pathways implicated in depression that were distinct to cancer survivors. Finally, we will discuss inflammation as a shared mechanism across cohorts in the context of previous hypotheses of cancer-associated depression.

### 3.4.1 Demographic and Clinical Characteristics

A slightly elevated percentage of cancer survivors in this study had high depressive symptoms compared to non-cancer participants (11.8 vs. 9.7%), relatively similar to previous US-based estimates.^1^ Several studies have previously reported the disproportionate impact of depression among active cancer patients,^50,51^ although consistent with our findings, these differences are often less pronounced in survivor populations.^52^

Furthermore, BMI was associated with depressive symptoms among cancer survivors but not non-cancer participants, although post-hoc comparisons showed only near-significant trends. Higher BMI is a well-established risk factor of depression in both cancer and general populations,^53,54^ although it remains unclear whether there may be distinct interactions between metabolism, inflammation, and depressive symptomology that are uniquely influenced by cancer and its treatments.^55,56^ Future research, potentially using metabolomic approaches, should investigate these interactions within cancer populations to provide further insight. Interestingly, we saw no differences between low and high depressive symptom groups in either cohort for age, gender, race/ethnicity, household income, and comorbidities that have previously been implicated as risk factors in large, population-based studies.^1,57,58^ This may be reflective of our smaller sample size or the distinct multi-ethnic diversity of MESA study participants relative to previous studies.

### 3.4.2 Neuronal Functioning, Neurotransmission, and Reward Circuitry

Common perturbations in depression across both cancer survivor and non-cancer cohorts included pathways involved in neuronal functioning and neurotransmission (e.g. neuroactive ligand-receptor interaction and calcium signaling), which are well-established in the pathophysiology of depression and support the validity of the pathway analysis.^11,59^ Notably, several neurodegenerative disease pathways (e.g. Alzheimer’s disease, Amyotrophic lateral sclerosis, Huntington’s disease) were distinctly perturbed in depression among cancer survivors. These pathways overlap in multiple biological processes previously implicated in the neurodegenerative hypothesis of depression.^60^ Specifically, increasing levels of oxidative stress inducing mitochondrial dysfunction and neuronal apotosis, which are hallmarks of the aging brain, have been linked with depression in preclinical models.^61^ Interestingly, cancer and anti-cancer therapies have been shown to disrupt these processes, and emerging evidence suggests that cancer patients exhibit accelerated brain aging linked with cognitive deficits.^62,63^ However, whether this accelerated brain aging plays a role in depressive symptoms specifically remains underexplored. The present findings suggest that future investigations of the interplay between accelerated aging pathways, specific therapies, and depression among cancer survivors are warranted, which can help inform the development of prevention and targeted treatment strategies.

Furthermore, several pathways related to dopaminergic signaling and reward circuitry (e.g. dopaminergic synapse, drug addiction and alcoholism pathways) were distinctly perturbed among cancer survivors with depression. Deficits in the dopaminergic system are strongly implicated in depression, culminating in core symptoms such as loss of interest or pleasure (i.e., anhedonia).^64,65^ These pathways being implicated only among cancer survivors in our study may reflect their exposure to cancer-specific biological and psychological stressors associated with dopaminergic dysregulation. For instance, cytotoxic chemotherapies commonly received by cancer patients have been shown *in vivo* to impair dopamine release in the striatum, a brain region essential for reward processing.^66,67^ It remains unclear whether these effects are short-term or may persist into survivorship. Furthermore, chronic psychosocial stress, which is reported by almost twice as many cancer survivors relative to the general population,^68^ dampens striatal dopaminergic function.^69^ Enhanced dopaminergic dysregulation may therefore play a role in the elevated prevalence of depression among cancer populations. Antidepressant strategies targeting dopaminergic and reward pathways (e.g. atypical antidepressants such as buproprion and mirtazapine) may be particularly applicable among cancer survivors, especially given their ability to modulate co-occuring symptoms such as insomnia, fatigue, and loss of appetite that may be contributing to depression. Future clinical studies are essential to understand their efficacy in this population.

Beyond pharmacological approaches, several of the pathways distinctly perturbed among cancer survivors, including dopaminergic signaling and circadian entrainment, correspond to mechanisms targeted by established psychosocial and behavioral interventions for depression. Behavioral activation therapy, an approach aimed at helping patients re-engage in meaningful activities, is proposed to function by targeting deficits in dopaminergic reward processing.^70^ Social rhythm therapy focuses on stabilizing daily routines such as the sleep/wake cycle to restore circadian function, with established efficacy for depression.^71^ Our findings provide mechanistic support for the potential utility of these interventions in cancer populations, warranting future clinical studies.

### 3.4.3 Cell Cycle, Growth, and Proliferation

Additionally, pathways related to cell cycle, growth, and proliferation were distinctly perturbed in depression among cancer survivors, including ErbB signaling and Hippo signaling. While these are largely known to be involved in cancer-related signaling, they also play important roles in synaptic plasticity and long-term potentiation by mediating neuronal differentiation, axonal growth, and cell apoptosis.^72,73^ Chronic stress dysregulates Hippo/YAP and neuregulin-ErbB signaling in pre-clinical models of depression,^74,75^ further implicating the role of population-specific experiences of psychological distress in shaping the biology underlying depression among cancer survivors. Given that it has also been hypothesized that the high prevalence of cancer-depression comorbidity implicates shared biological perturbations,^2,20–22^ further investigation of these pathways can elucidate whether overlaps in cancer-related and neuronal proliferative signaling may drive depressive symptoms in cancer populations. Finally, distinct perturbations of oocyte meiosis and progesterone-mediated oocyte maturation pathways may implicate a role for dysregulated hormone signaling in depression among cancer survivors, potentially relating to an overactive HPA axis or exposure to endocrine therapies.^2^ While our cohort included both male and female cancer survivors, future studies with sample sizes sufficient for stratified analyses could explore whether these perturbations may be sex-specific.

### 3.4.4 Metabolism, Endocrine Signaling, and Cardiovascular Pathways

In addition to neuronal and proliferative pathways, three secretory pathways (salivary, gastric acid, and pancreatic secretion) were perturbed in depression uniquely among cancer survivors. Little is known about the role of these pathways in depression, although they may represent the dysregulation of the autonomic nervous system and neuroendocrine function, including HPA axis overactivity, that has been observed in clinical studies of major depressive disorder.^76–78^ Furthermore, cytotoxic chemotherapies damage mucosal epithelial cells in gastrointestinal and salivary systems, leading to tight junction and secretory dysfunction.^79,80^ The resulting side effects of oral and gastrointestinal mucositis are prevalent in up to 100% of patients receiving these treatments and are highly correlated with depressive symptoms.^81–83^ Our findings support further evaluation of stress and treatment-induced dysregulation of autonomic and neuroendocrine systems, and whether these effects may persist into survivorship, as potential mechanisms of depression among cancer survivors.

Additionally, the perturbation of cardiovascular pathways may further implicate autonomic dysregulation in depression among cancer survivors.^76,78^ However, aside from the well-known prevalence of comorbidities of cancer, cardiovascular disease, and depression,^84^ how these pathways may overlap to drive these conditions remains poorly understood, warranting further investigation.

### 3.4.5 Inflammatory Processes and Immune Signaling

Finally, one of the predominant theories of cancer-associated depression involves the dysregulation of inflammatory processes and immune signaling.^2,10,14,49^ Preclinical and clinical studies demonstrate that cancer and anti-cancer therapies can induce peripheral inflammation, which can contribute to neuroinflammation and oxidative stress and culminate in neuropsychiatric symptoms such as depression.^12,14^ While this mechanistic category had the greatest number of perturbed pathways in the present study, notably, 28 out of the 29 inflammatory pathways implicated in depression among cancer survivors were also perturbed in the non-cancer cohort. While this does not preclude the potential for these pathways to be perturbed in different ways or to a greater extent, it does suggest that inflammation represents a common mechanism of depression among both cancer survivors and non-cancer populations. Consistent with the general population, targeting inflammation may serve as a potential therapeutic strategy to address depression among cancer survivors.^13^

The sole perturbed inflammatory pathway that was distinct to cancer survivors was Fc gamma R-mediated phagocytosis. In the brain, microglia utilize this process to clear debris, and defects in microglial phagocytosis have been linked to depression.^85^ While the role of Fc gamma R-mediated phagocytosis in cancer-associated depression warrants further investigation, our findings strongly support inflammatory dysregulation as a whole as a shared mechanism of depression across cancer and non-cancer groups.

### 3.4.6 Limitations and Future Directions

While our findings provide important insights on mechanisms underlying depressive symptoms among cancer survivors, several limitations must be noted. First, it is important to note that the CES-D is a screening tool, and the cutoff represents those considered at risk for clinical depression rather than diagnosed cases of MDD. Second, given the cross-sectional design of this study, causal relationships between pathway perturbations and depressive symptoms cannot be ascertained. Future clinical and preclinical studies are warranted to investigate whether these pathways are direct contributors to depression and to evaluate the efficacy of therapeutics targeting these pathways. Third, due to the availability of non-cancer controls, matching to cancer survivors was based on age alone. Future studies with larger sample sizes should consider additional matching covariates to further isolate the effect of cancer survivorship status. Finally, detailed data on time from cancer diagnosis and treatment history were unavailable for the current study. Future studies evaluating how perturbed pathways associated with depression differ among individual cancer types, stages, and treatment regimens may provide further mechanistic insight.

### 3.5 Conclusions

In conclusion, our findings implicate a unique role of mechanisms related to neurodegeneration, reward circuitry, autonomic secretion, and distinct cell cycle and proliferative processes in depression among cancer survivors, suggesting that cancer-specific biological and psychological stressors may shape a distinct pathophysiology underlying these debilitating symptoms. Notably, they also implicate inflammation as a shared mechanism of depression across cancer and non-cancer populations. Given that depression represents a major unmet need in cancer survivorship care, our findings provide strong support for the use of precision medicine approaches and identify therapeutic targets that can be prioritized in the evaluation of novel or repurposed interventions.

## Author Contributions

**Julia E. Trudeau**: conceptualization, data curation, formal analysis, investigation, methodology, visualization, writing – original draft, writing – review and editing. **Nidhi Thati**: investigation, writing – review and editing. **Ding Quan Ng**: writing – review and editing. **Esther Chavez-Iglesias**: writing – review and editing. **Adam B. Olshen**: methodology, writing – review and editing. **Anand Dhruva**: writing – review and editing. **Jason W. Chan**: writing – review and editing. **Raymond J. Chan**: conceptualization, writing – review and editing. **Alexandre Chan**: conceptualization, funding acquisition, methodology, writing – review and editing. **Kord M. Kober**: conceptualization, data curation, funding acquisition, formal analysis, methodology, project administration, resources, software, supervision, writing – original draft, writing – review and editing.

## Acknowledgements

Research reported in this study was supported by grants from the National Cancer Institute (Kober: R37CA233774, Chan: R01CA276212) and a Cancer Center Support Grant (P30, CA082103). Its contents are solely the responsibility of the authors and do not necessarily represent the official views of the NIH. The Multi-Ethnic Study of Atherosclerosis (MESA) study was supported by the National Heart, Lung, and Blood Institute (contracts 75N92020D00001, HHSN268201500003I, N01-HC-95159, 75N92020D00005, N01-HC-95160, 75N92020D00002, N01-HC-95161, 75N92020D00003, N01-HC-95162, 75N92020D00006, N01-HC-95163, 75N92020D00004, N01-HC-95164, 75N92020D00007, N01-HC-95165, N01-HC-95166, N01-HC-95167, N01-HC-95168, N01-HC-95169); and the National Center for Advancing Translational Sciences (grants UL1-TR-000040, UL1-TR-001079, UL1-TR-001420). We gratefully acknowledge the MESA study investigators, staff, and participants for their contributions, without whom this research would not have been possible.

## Ethical approval statement

De-identified data from the MESA study were obtained through an approved study project (39341) through the National Institutes of Health (NIH) database of Genotypes and Phenotypes (dbGaP). The MESA study was approved by institutional review boards at all participating study sites, and all participants provided informed consent.

## Data availability statement

Data used in this study are available for qualified researchers through dbGaP under accession numbers phs000209.v13.p3 (Multi-Ethnic Study of Atherosclerosis (MESA) Cohort) and phs001416.v4.p1 (NHLBI TOPMed MESA and MESA Family AA-CAC).

