## Supplementary Material for "Distinct neuronal, proliferative, and secretory pathways are perturbed in cancer survivors with depressive symptoms"

**This PDF file includes:**

Supplementary Table 1. Center for Epidemiologic Studies Depression Scale (CESD-20) items given to MESA Study participants.

Supplementary Table 2. Sociodemographic characteristics of cancer survivors and non-cancer cohorts before and after age-matching.

Supplementary Figure 1. Participant selection flowchart.

**Supplemental Data Tables (in separate documents):**

Supplemental File 1. Pathway impact analysis for cancer survivors.

Supplemental File 2. Pathway impact analysis for non-cancer cohort.

Supplemental File 3. Differential expression analysis for cancer survivors.

Supplemental File 4. Differential expression analysis for non-cancer cohort.

Supplementary Table 1. Center for Epidemiologic Studies Depression Scale (CESD-20) items given to MESA Study participants. CESD-20 items are self-reported and measure depressive symptoms over the past week.

| Item | Below is a list of the ways you might have felt or behaved. Please indicate how often you felt this way DURING THE PAST WEEK.[a] |
|---|---|
| A | I was bothered by things that don't usually bother me |
| B | I did not feel like eating; my appetite was poor |
| C | I felt that I could not shake off the blues, even with help from my family and friends |
| D | I felt that I was just as good as other people |
| E | I had trouble keeping my mind on what I was doing |
| F | I felt depressed |
| G | I felt that everything I did was an effort |
| H | I felt hopeful about the future |
| I | I thought my life had been a failure |
| J | I felt fearful |
| K | My sleep was restless |
| L | I was happy |
| M | I talked less than usual |
| N | I felt lonely |
| O | People were unfriendly |
| P | I enjoyed life |
| Q | I had crying spells |
| R | I felt sad |
| S | I felt that people dislike me |
| T | I could not "get going" |

[a] Responses are scored on a 4-point scale: 0 = Rarely or none of the time (<1 day), 1 = Some or a little of the time (1–2 days), 2 = A moderate amount of the time (3–4 days), 3 = Most of the time (5–7 days). Total scores range from 0–60, with higher scores indicating more depressive symptoms. An established cutoff of ≥16 is used to indicate a person at risk of clinical depression.

Supplementary Table 2. Sociodemographic characteristics of cancer survivors and non-cancer cohorts before and after age-matching.

| Characteristic | Cancer survivors (*n*=93) | Unmatched (whole) Non-cancer Cohort (*n*=1045) | Statistic | Age-matched Non-cancer Cohort (*n*=176) | Statistic |
|---|---|---|---|---|---|
| | % (n) | % (n) | | % (n) | |
| Age | | | | | |
| 45-54 | 3% (3) | 36% (372) | | 3% (6) | |
| 55-64 | 15% (14) | 30% (315) | $x^2$=168.67, p<0.001 | 16% (28) | $x^2$=0.19, p=0.975 |
| 65-74 | 31% (29) | 26% (274) | | 33% (58) | |
| 75-84 | 51% (47) | 8% (84) | | 48% (84) | |
| Gender | | | | | |
| Female | 52% (48) | 54% (564) | OR=1.10, p=0.666 | 57% (100) | OR=1.23, p=0.441 |
| Male | 48% (45) | 46% (481) | | 43% (76) | |
| Race/Ethnicity | | | | | |
| White, Caucasian | 67% (62) | 35% (361) | | 34% (59) | |
| Chinese American | 6% (6) | 10% (103) | $x^2$=47.60, p<0.001 | 14% (24) | $x^2$=36.37, p<0.001 |
| Black, African-American | 26% (24) | 29% (305) | | 31% (54) | |
| Hispanic | 1% (1) | 26% (276) | | 22% (39) | |
| Household income | | | | | |
| <$29,999 | 34% (32) | 34% (351) | | 50% (88) | |
| $30,000 - $74,999 | 40% (37) | 46% (479) | $x^2$=3.29, p=0.353 | 38% (67) | $x^2$=10.50, p=0.017 |
| $75,000 - $100,000 | 9% (8) | 9% (97) | | 3% (6) | |
| >$100,000 | 17% (16) | 11% (118) | | 9% (15) | |
| Marital status | | | | | |
| Not married | 41% (38) | 39% (409) | OR=0.93, p=0.741 | 47% (83) | OR=1.29, p=0.368 |
| Married | 59% (55) | 61% (636) | | 53% (93) | |
| BMI | | | | | |
| Normal | 30% (28) | 24% (253) | | 29% (51) | |
| Grade 1 Overweight | 40% (37) | 38% (401) | $x^2$=2.49, p=0.473 | 38% (67) | $x^2$=3.57, p=0.330 |
| Grade 2 Overweight | 27% (25) | 33% (347) | | 32% (57) | |
| Grade 3 Overweight | 3% (3) | 4% (44) | | 1% (1) | |

Abbreviations: BMI = Body mass index

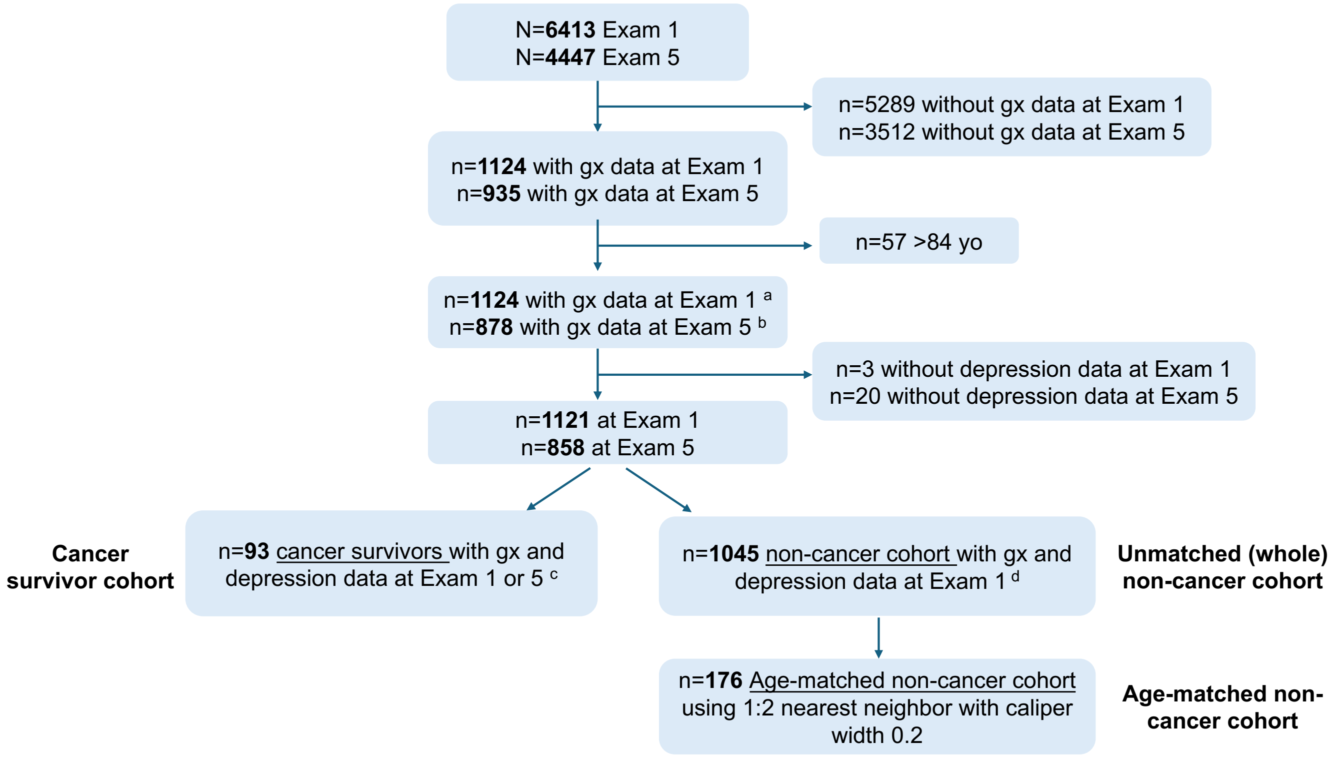


Supplementary Figure 1. Participant selection flowchart. 93 cancer survivors with depression and gene expression data were analyzed. A non-cancer cohort (n=176) was age-matched to the cancer survivors.

[a] Multiple imputation was performed for missing covariate data as follows (all <5% missingness): Household income (n=17), Marital status (n=12), Antidepressant use (n=1), Diabetes (n=1).
[b] Multiple imputation was performed for missing covariate data as follows (all <5% missingness): Household income (n=26), Marital status (n=24), Antidepressant use (n=3), Diabetes (n=1), Hypertension (n=1).
[c] For cancer survivors with gene expression data available at both exams, only one observation was selected to maintain independence. Exam 5 was selected first and backfilled with Exam 1 if not available.
[d] The non-cancer cohort were selected from Exam 1 given that diagnosis history was not re-assessed at Exam 5 to avoid misclassification of newly diagnosed survivors.

Abbreviations: gx=Gene expression
